# Clinical and serological findings of Madariaga and Venezuelan equine encephalitis viral infections: A follow-up study five years after an outbreak in Panama

**DOI:** 10.1101/2020.02.13.20022798

**Authors:** Jean-Paul Carrera, Yaneth Pittí, Juan C. Molares-Matrínez, Eric Casal, Reneé Pereyra-Elias, Lisseth Saenz, Isela Guerrero, Josefrancisco Galué, Fatima Rodriguez-Alvarez, Carmela Jackman, Juan Miguel Pascale, Blas Armien, Scott C. Weaver, Christl A. Donnelly, Amy Y. Vittor

**Affiliations:** Department of Zoology, University of Oxford, Oxford, United Kingdom; Department of Research in Virology and Biotechnology, Gorgas Memorial Institute of Health Studies, Panama City, Panama; National Perinatal Epidemiology Unit, Nuffield Department of Population Health, University of Oxford, Oxford, United Kingdom; Departments of Epidemiology, Ministry of Health, Panama, Panama; Clinical Research Unit, Gorgas Memorial Institute of health studies, Panama City, Panama; Department of Research in Emerging and Zoonotic Diseases, Gorgas Memorial Institute of Health Studies, Panama City, Panama; Institute for Human Infection and Immunity, Department of Microbiology and Immunology, Department of Pathology, and World Reference Center for Emerging Viruses and Arboviruses, University of Texas Medical Branch, Galveston, Texas; MRC Centre for Global Infectious Disease Analysis, Department of Infectious Disease Epidemiology, Imperial College London, London, United Kingdom; Department of Statistics, University of Oxford, Oxford, United Kingdom; Department of Medicine, Division of Infectious Disease and Global Medicine, University of Florida, Gainesville, Florida

**Keywords:** Madariaga virus, Venezuelan equine encephalitis virus, neurological sequelae, Panama

## Abstract

**Background:** Human cases of Madariaga virus (MADV) infection were first detected during an outbreak in 2010 in eastern Panama, where Venezuelan equine encephalitis virus (VEEV) also circulates. Little is known about the long-term consequences of either alphavirus infection.

**Methods:** A follow-up study of the 2010 outbreak was undertaken in 2015. An additional survey was carried out two weeks after a separate 2017 alphavirus outbreak in a neighboring population in eastern Panama. Serological studies and statistical analysis were undertaken in both populations.

**Results:** Amongst the originally alphavirus-seronegative subjects (n=35 of 65), seroconversion was observed at a rate of 14.3% (95% CI: 4.8%-30.3%) for MADV and 8.6% (95% CI: 1.8%-23.1%) for VEEV over 5 years. Amongst the originally MADV seropositive subjects (n=14 of 65), VEEV seroconversion occurred in 35.7% (95% CI: 12.8%-64.9%). In the VEEV seropositive subjects (n=16 of 65), MADV seroconversion occurred in 6.3% (95% CI: 0.2%-30.2%). MADV seroreversion was observed in 14.3% (95% CI: 1.8%-42.8%) of those originally seropositive in 2010. VEEV seroconversion in the baseline MADV-seropositive subjects was significantly higher than in alphavirus-negative subjects. In the population sampled in 2017, MADV and VEEV seroprevalence was 13.2% and 16.8%, respectively. Memory loss, insomnia, irritability and seizures were reported significantly more frequently in alphavirus-seropositive subjects than in seronegative.

**Conclusions:** High rates of 5-year seroconversions to MADV and VEEV suggest continuous circulation of both viruses in Panama. Enhanced susceptibility may be conferred by MADV towards VEEV. We provide evidence of persistent neurologic symptoms up to 5 years following MADV and VEEV exposure.

**summary:** We estimate seroconversion rates over a 5-year period to Madariaga (MADV) and Venezuelan equine encephalitis (VEEV) alphaviruses in Panama. Individuals with MADV antibodies seroconverted to VEEV at a rate greater than individuals who were alphavirus-negative at baseline. This was not observed in individuals with VEEV antibodies, suggesting asymmetric cross-immunity. Neurological sequelae were reported more frequently by MADV and/or VEEV seropositive-versus seronegative subjects.

## Introduction

Madariaga (MADV, formerly known as South American eastern equine encephalitis) and Venezuelan equine encephalitis viruses (VEEV) are single stranded RNA arthropod-borne zoonotic viruses (Togaviridae: Alphavirus), with circulation throughout much of the Americas [1]. Enzootic subtypes of the VEE antigenic complex are associated with human endemic, and sometimes fatal, infections in the Americas [2]. Human infections with these subtypes occur via spillover from enzootic cycles that involve sylvatic rodents and mosquitoes of the subgenus Culex (Melanoconion). VEEV epizootic/epidemic subtypes (IAB, IC) are associated with large and explosive equine-amplified epidemics in South America and the available evidence suggest that epizootics strains evolve from enzootic ancestors via mosquito or equine-adaptive mutations [3]. In Panama, enzootic/endemic VEEV subtype ID infection is highly prevalent in the easternmost province of Darien, resulting in up to 75% seroprevalence in some villages [4]. On this background, MADV first emerged in the human population in 2010 in the eastern province of Darien, Panama [5]. Enzootic VEEV (subtype ID) was simultaneously circulating in the same area, causing significant neurologic morbidity and mortality [5].

Clinically, most human VEEV infections are symptomatic with dengue or flu-like illness [6]. VEE is under-diagnosed in Latin America, where it has been estimated that 0.1-7% of dengue cases are in fact VEEV infections [2]. Around 15% of VEEV cases develop neurologic disease, of which 1% are fatal [2]. In contrast, only 3 human cases of MADV were identified in the Americas prior to the 2010 outbreak in Panama, despite extensive research and epidemiologic surveillance in enzootic areas [7–9]. Mosquito vectors of MADV are also members of the subgenus Culex (Melanoconion). However, the main reservoir in Latin America is still unknown[10].

The acute clinical presentation of MADV and VEEV infections has been described during outbreaks or sporadic cases [5,11,12]. However, because MADV is an emerging virus in Latin America, the long-term sequelae remain unknown. Our literature search revealed little information about long-term neurologic sequelae of enzootic VEEV infections. Following an outbreak in Texas in 1971, Bowen et al. (1976) described signs and symptoms in 86 patients hospitalized with VEEV[6]. None of the affected children reported any sequelae, but seven of nine adults examined 9 months later complained of fatigue.

Here we characterize the clinical and serological consequences of MADV and VEEV infections by following up on probable and confirmed cases and their household contacts identified during the 2010 outbreak. We also validate the clinical features of the 2010 cohort by including results of the neurological sign/symptom survey of a neighboring population in Darien province.

## Methods

The study was undertaken in the easternmost province of Darien, Panama, which borders Colombia. Alphaviral encephalitis outbreaks were reported in Darien in 2010 and 2017. A follow-up study of patients (suspected, probable or confirmed) from the 2010 outbreak and their household contacts was undertaken in 2015. Detailed information on the 2010-2015 cohorts is provided in Figure 1 and Table 1. An additional population survey was undertaken in 2017 in Mogue, a community in Darien that was not included in the passive surveillance efforts of 2010.

**Table 1.**
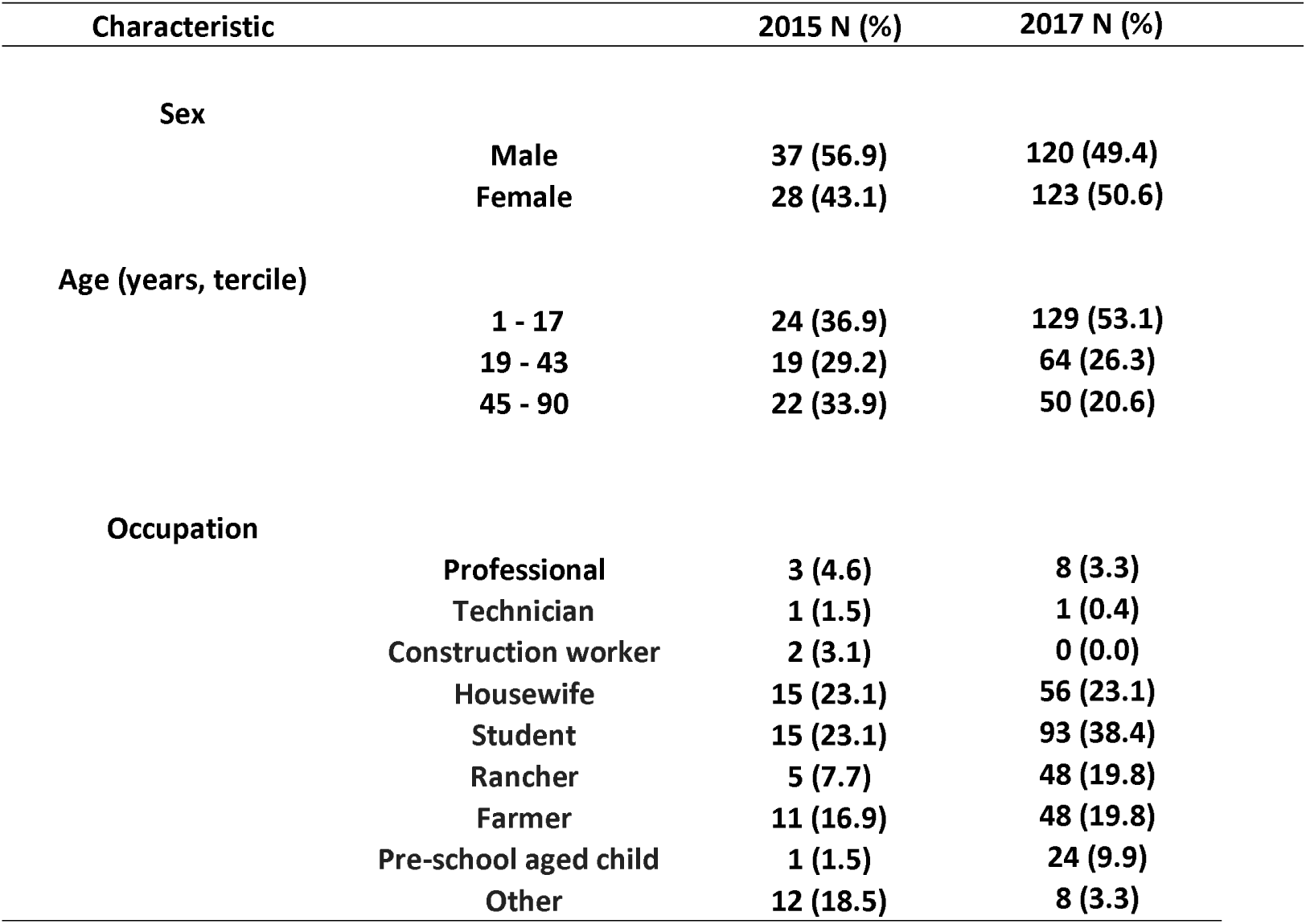
Characteristics of the study populations in the 2010 outbreak cohort studied in 2015, n=65, and the Mogue study in 2017, n= 243.

| Characteristic |  | 2015 N (%) | 2017 N (%) |
| --- | --- | --- | --- |
| **Sex** |  |  |  |
|  | **Male** | **37 (56.9)** | **120 (49.4)** |
|  | **Female** | **28 (43.1)** | **123 (50.6)** |
| **Age (years, tercile)** |  |  |  |
|  | **1 - 17** | **24 (36.9)** | **129 (53.1)** |
|  | **19 - 43** | **19 (29.2)** | **64 (26.3)** |
|  | **45 - 90** | **22 (33.9)** | **50 (20.6)** |
| **Occupation** |  |  |  |
|  | **Professional** | **3 (4.6)** | **8 (3.3)** |
|  | **Technician** | **1 (1.5)** | **1 (0.4)** |
|  | **Construction worker** | **2 (3.1)** | **0 (0.0)** |
|  | **Housewife** | **15 (23.1)** | **56 (23.1)** |
|  | **Student** | **15 (23.1)** | **93 (38.4)** |
|  | **Rancher** | **5 (7.7)** | **48 (19.8)** |
|  | **Farmer** | **11 (16.9)** | **48 (19.8)** |
|  | **Pre-school aged child** | **1 (1.5)** | **24 (9.9)** |
|  | **Other** | **12 (18.5)** | **8 (3.3)** |

**Table 2.**
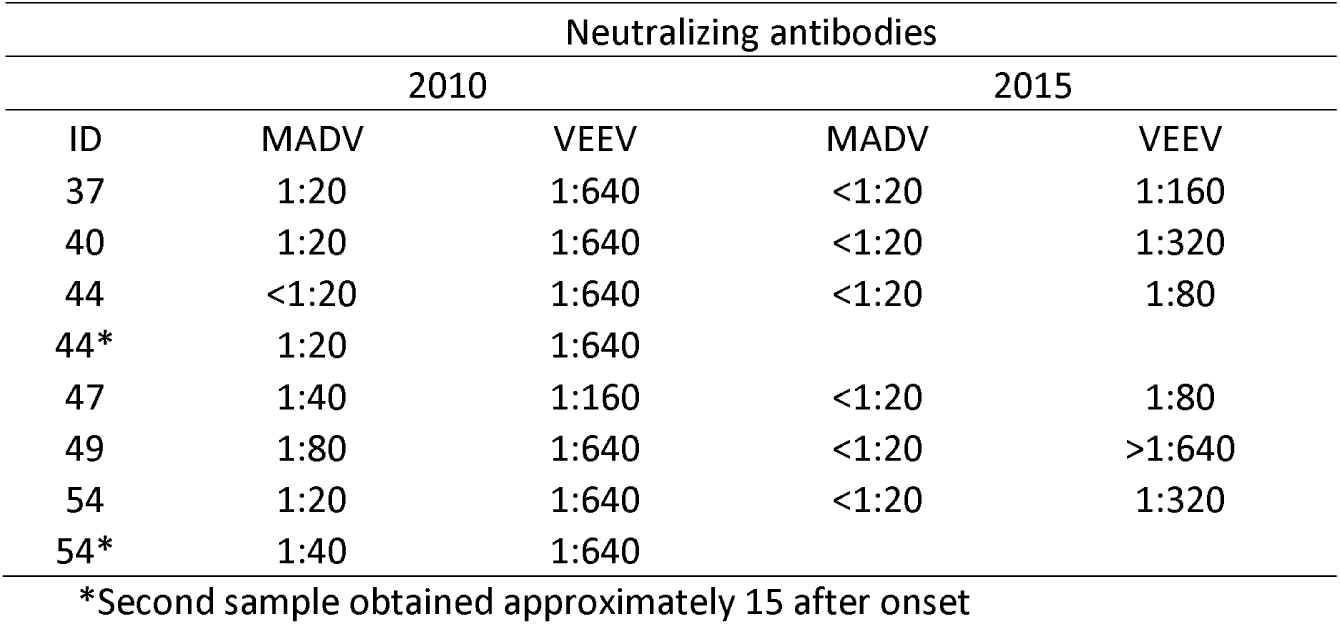
Neutralizing antibody titters measured in 2010 and 2015 among the 6 individuals found to have antibodies to both MADV and VEEV in 2010.

**Figure 1.**
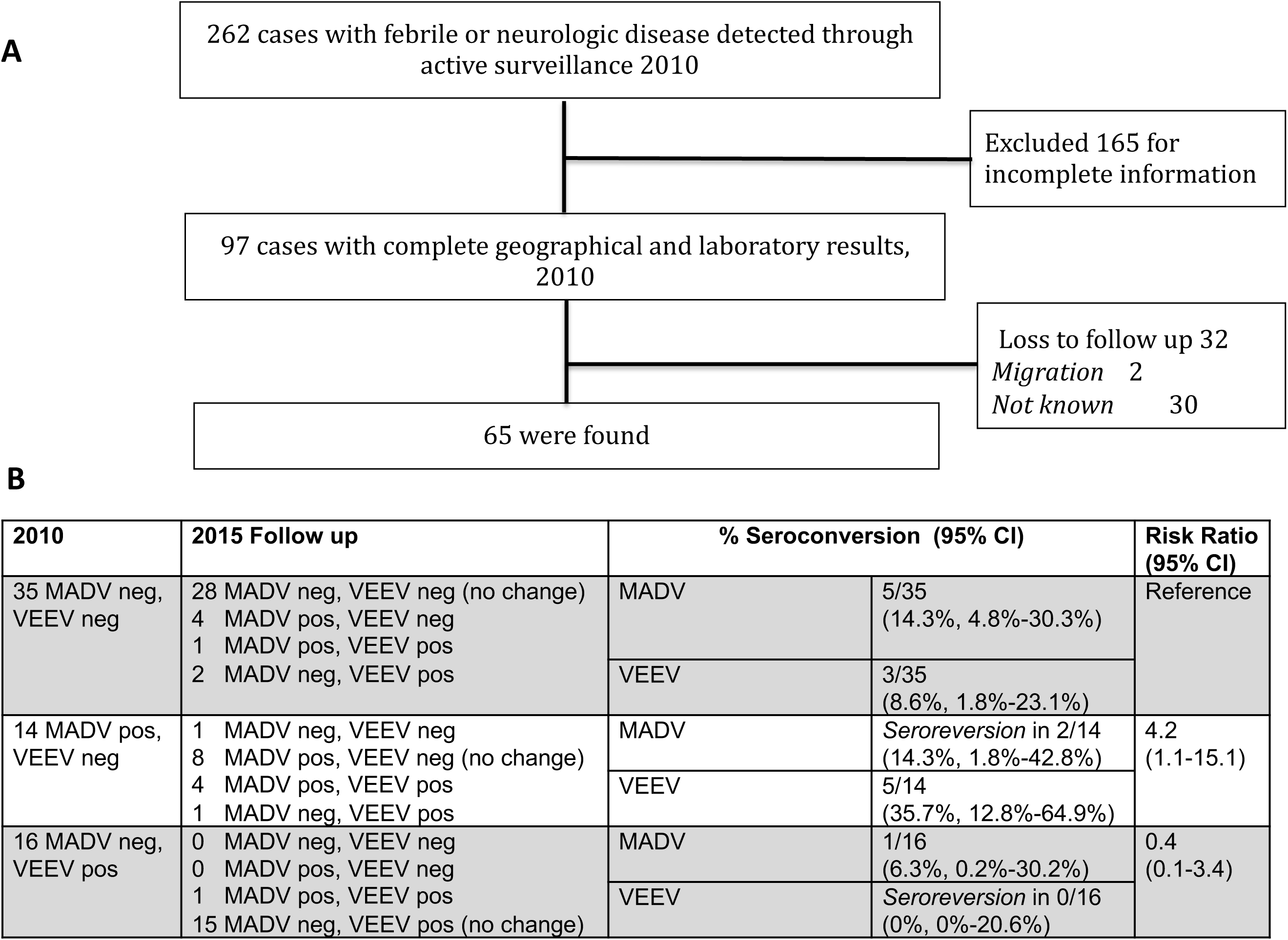
Epidemiological flowchart and seroconversion in the 2010 original outbreak cohort. **A)** Description of the criteria used for selection of the. **B)** Risk ratios compare the probability of seroconversion to heterologous virus in the MADV or VEEV exposed group versus the probability of seroconversion in the alphavirus seronegative group.

### Human survey and clinical evaluation in 2015

Subjects were located using records from 2010. Upon obtaining written consent, each participant was interviewed and examined by a physician using a demographic and focused medical history questionnaire. The presence or absence of neurological symptoms within the preceding 2 weeks was noted. Blood samples were obtained by peripheral venipuncture. Serum samples were placed in cryogenic tubes for storage in liquid nitrogen and then transported to the Gorgas Memorial Institute for analysis.

### Human survey and clinical evaluation in 2017

The survey was carried out from July 18-22, 2017, three weeks after confirmation of a fatal MADV case on June 30, 2017. A cross-sectional outbreak investigation was undertaken, and blood sampling and epidemiological surveys were carried out including demographic characteristics, potential risk factors and clinical information. The presence or absence of neurological symptoms within the preceding 2 weeks was evaluated and recorded by a physician. Detailed information on surveys, laboratory testing, risk factors and serologic results are were previously described [13]. The purpose of this 2017 survey was to identify additional cases of severe encephalitis and neurological sequelae in a community where a MADV fatal case was confirmed. We include the 2017 neurological symptom survey here to validate the findings from our 2010 cohort.

### Ethics

Ethic approval was for alphaviruses encephalitis surveillance and investigation was obtained from the Gorgas Memorial Institute Committee (IRB, CBI/ICGES/2015 and CIB/ICGES/2018). Study participation was voluntary, and written informed consent was obtained from adults (18 years of age and older) and from parents or guardians for children aged 2 to 17 years. In addition, verbal assent was obtained from children aged 7 to 12 years, and written informed consent was obtained from children aged 13 to 17 years.

### Laboratory

Samples were also tested in duplicate for IgM and IgG antibodies against MADV and VEEV using an enzyme-linked immunosorbent assays (ELISA) for recent and past antibody response detection [14]. All samples were also run using virus-specific plaque reduction neutralization tests (PRNTs). For the PRNT, a positive sample was reported as the reciprocal of the highest dilution that reduced plaque counts by >80% (PRNT_80_). ELISA antigens were prepared from EEEV by the sucrose-acetone technique (prepared by Dr. Robert Shope at the Yale Arbovirus Research Unit in August 1989) and VEE complex strain 78V-3531 from infected mouse brain. Strains used for the PRNT were the chimeric SINV/MADV [15], as well as MADV wild-type strain PA2010-247168 (UTMB/WRCEVA) and TC83, an attenuated vaccine strain of VEEV closely related to subtype ID strains that circulate in Panama [16].

### Statistical analysis

Seroconversion rates for both viruses were calculated for each alphaviral exposure group from the 2010 cohort and reported with exact binomial 95% confidence intervals. The probability of seroconversion by alphaviral exposure group was determined by calculating risk ratios with exact Fisher’s exact p given the small sample sizes. To determine whether neurological sequelae were reported at higher frequencies in the alphavirus-exposed group, we conducted a univariate logistic regression analysis, followed by a multivariable logistic regression controlling for sex and age. The outcome variable, alphaviral exposure, was defined as having been exposed to MADV, VEEV, or both viruses using the laboratory criteria described above. P-values with alpha < 0.05 were considered to be significant. All analyses were undertaken using the statistical package Stata v14 (StataCorp, College Station, TX, 2014).

## Results

### Characteristics of the population

During the original 2010 outbreak, 190 cases with febrile or neuroinvasive disease and 72 household contacts were included. In the 2015 follow-up study, 165 of these subjects were excluded due to incomplete information and an additional 32 cases could not be located. A total of 65 cases were ultimately included in the 2015 follow-up serosurvey (Figure 1. A.). Characteristics of the 2015 and Mogue 2017 study populations are described in Table 1.

### Alphavirus epidemiological and serological profiles

#### Study design

We undertook two separate efforts to survey human population in Darien province: 1) The first was a 2015 follow-up study of the original 2010 cases; 2) An additional cross-sectional outbreak investigation was used to validate the clinical outcomes reported for the 2010 original cohort.

#### Baseline results

Using virus-specific PRNTs, a total of 14 (21.5%) of the 65 subjects in the follow-up study were seropositive for MADV only, 16 (24.6%) were seropositive for VEEV only, and 0 were seropositive for both at baseline in 2010 (Figure 1. B).

#### Follow up in 2015

Amongst the 35 subjects who were alphavirus-seronegative in 2010, seroconversion was observed for MADV in 5 of 35 (14.3%, 95% CI: 4.8%-30.3%) and VEEV in 3 of 35 (8.6%, 95% CI: 1.8%-23.1%) by 2015 (Figure 1. B). Amongst the 14 subjects who were MADV-seropositive in 2010, seroconversion to VEEV was observed in 2015 in 5 of 14 (35.7%, 95% CI: 12.8%-64.9%). Only one of the 16 subjects who were VEEV-seropositive in 2010 seroconverted to MADV by 2015 (6.3%, 95%CI: 0.2%-30.2%). Seroreversion (disappearance of detectable antibodies) was observed for MADV in 2 of 14 subjects (14.3%, 95%CI: 1.8%-42.8%). No VEEV seroreversions were observed (0%, 95% CI: 0%-20.6%).

The risk ratio for seroconversion to MADV in VEEV-seropositive (in 2010) subjects versus previously MADV- and VEEV-seronegative subjects was 0.4 (95% CI: 0.1-3.4; Fisher’s exact p=0.38). The risk ratio for seroconversion to VEEV in MADV-seropositive (in 2010) subjects versus MADV- and VEEV-seronegative subjects was 4.2 (95% CI: 1.1-15.1; Fisher’s exact p=0.03).

A total of 6 individuals were found to have neutralizing antibodies to both MADV and VEEV in 2010, though in each instance titers were at least 4-fold greater against VEEV (Table 3). Each of these individuals with low MADV titers became MADV PRNT_80_-negative (≥1:20) by 2015 (Figure 1. B).

**Table 3.** Frequency of neurological signs and symptoms (%) reported in two study populations in Darien, Panama. Exposure status reflects seropositivity measured in 2015 for the original outbreak cohort, and seropositivity measured in 2017 for the Mogue study.

| *Symptoms* | *Original Outbreak Cohort (n=65)* |  |  |  | *2017 Mogue Study (n=243)* |  |  |  |
| --- | --- | --- | --- | --- | --- | --- | --- | --- |
|  | *MADV*<br>*(n=13)* | *VEEV*<br>*(n=17)* | *MADV &*<br>*VEEV*<br>*(n=7)* | *No*<br>*Alphaviral*<br>*exposure*<br>*(n=28)* | *MADV*<br>*(n=22)* | *VEEV*<br>*(n=33)* | *MADV &*<br>*VEEV*<br>*(n=9)* | *No*<br>*Alphaviral*<br>*exposure*<br>*(n=179)* |
| *Memory loss* | 6 (46) | 8 (47) | 4 (57) | 8 (29) | 8 (36) | 10 (30) | 3 (33) | 37 (21) |
| *Headache* | 5 (42)* | 7 (41) | 4 (57) | 11 (39) | 9 (41) | 15 (45) | 6 (67) | 80 (45) |
| *Dizziness* | 5 (38) | 6 (35) | 4 (57) | 5 (18) | 7 (32) | 13 (39) | 5 (56) | 47 (26) |
| *Fatigue* | 4 (31) | 5 (29) | 3 (43) | 4 (14) | 10 (45) | 14 (42) | 5 (56) | 56 (31) |
| *Confusion* | 5 (38) | 5 (29) | 1 (14) | 2 (7) | 3 (14) | 8 (24) | 3 (33) | 27 (15) |
| *Depression* | 2 (17) | 5 (29) | 3 (43) | 3 (11) | 5 (23) | 1 (3) | 1 (11) | 15 (8) |
| *Irritability* | 4 (31) | 5 (29) | 1 (14) | 1 (4) | 1 (5) | 3 (9) | 1 (11) | 11 (6) |
| *Myalgia* | 2 (17) | 3 (18) | 3 (43) | 4 (14) | 8 (36) | 9 (27) | 4 (44) | 40 (22) |
| *Insomnia* | 3 (23) | 4 (24) | 2 (29) | 3 (11) | 3 (14) | 6 (18) | 6 (67) | 18 (10) |
| *Difficulty with*<br>*activities of*<br>*daily living* | 3 (23) | 1 (6) | 1 (14) | 1 (4) | 6 (27) | 6 (18) | 2 (22) | 20 (11) |
| *Weakness* | 1 (8) | 3 (18) | 1 (14) | 2 (7) | 9 (41) | 9 (27) | 4 (44) | 43 (24) |
| *Seizures* | 2 (15) | 0 (0) | 0 (0) | 0 (0) | 2 (9) | 2 (6) | 0 (0) | 1 (0.6) |
| *Paralysis* | 1 (8) | 0 (0) | 0 (0) | 0 (0) | 1 (5) | 4 (12) | 0 (0) | 6 (3) |
| *Total* | 13 | 17 | 7 | 28 | 22 | 33 | 9 | 179 |

***N.B. subjects whose MADV titers seroreverted in 2015 are classified as MADV positive \* one non-respondent in this category yielded a denominator of 12***

#### Seroprevalence in 2017

A total of 243 individuals were surveyed in Mogue, Darien Province during 2017, with seroprevalence of 32/243 (13.2%) for MADV and 41/243 (16.8%) for VEEV.

#### Neurological symptoms

Memory loss, dizziness, fatigue, difficulty concentrating, confusion, depression, irritability, myalgia, insomnia, seizures, and impairment in activities of daily living were more frequent in subjects exposed to VEEV and/or MADV (Table 4, Figure 2). We repeated this survey in a different population in Darien, Panama, in 2017, with roughly similar results (Table 3, right-hand columns). After adjusting for sex and age, the association with prior alphaviral (MADV and/or VEEV) exposure and seizures, irritability, insomnia and memory loss remained statistically significant (Table 4).

**Table 4.** Univariate and multivariable logistic regression analysis of the presence of self-reported signs and symptoms by alphaviral exposure (n= 308).

| Symptoms | n (%) | Unadjusted<br>Odd Ratio | IC 95% | P value | φ Adjusted<br>Odds Ratio | IC 95% | P value |
| --- | --- | --- | --- | --- | --- | --- | --- |
| Seizures | 7 (2.3) | 13.21 | 1.56-111.3 | 0.018 | 14.46 | 1.61-130.1 | 0.017 |
| Paralysis | 12 (3.9) | 2.14 | 0.68-6.83 | 0.195 | 1.5 | 0.43-5.30 | 0.525 |
| Difficulty walking | 29 (9.4) | 1.10 | 0.49-2.47 | 0.808 | 0.87 | 0.36-2.10 | 0.767 |
| Irritability | 27(8.8) | 2.44 | 1.10-5.41 | 0.028 | 2.68 | 1.14-6.27 | 0.023 |
| Depression* | 35(11.4) | 1.92 | 0.94-3.91 | 0.074 | 1.73 | 0.81-3.65 | 0.154 |
| Impairment in ADL | 40 (13.0) | 2.08 | 1.06-4.09 | 0.032 | 1.47 | 0.71-3.03 | 0.297 |
| Insomnia | 45 (14.6) | 2.81 | 1.48-5.35 | 0.002 | 2.52 | 1.28-4.97 | 0.008 |
| Confusion | 54 (17.5) | 1.88 | 1.03-3.41 | 0.040 | 1.64 | 0.87-3.10 | 0.128 |
| Weakness | 72(23.4) | 1.34 | 0.77-2.32 | 0.298 | 1.18 | 0.66-2.11 | 0.564 |
| Myalgia | 73 (23.7) | 1.52 | 0.88-2.63 | 0.131 | 1.10 | 0.61-1.98 | 0.757 |
| Memory Loss | 84 (27.3) | 2.31 | 1.38-3.90 | 0.002 | 1.89 | 1.09-3.28 | 0.022 |
| Dizziness | 92(29.9) | 1.87 | 1.12-3.10 | 0.016 | 1.48 | 0.87-2.54 | 0.148 |
| Fatigue | 1001(32.8) | 1.61 | 0.98-2.65 | 0.063 | 1.27 | 0.74-2.16 | 0.376 |
| Headache | 137 (44.6) | 1.05 | 0.65-1.70 | 0.840 | 0.86 | 0.51-1.45 | 0.582 |

\*N=307
φAdjusted by age and sex: ADL= activities of daily living

**Figure 2.**
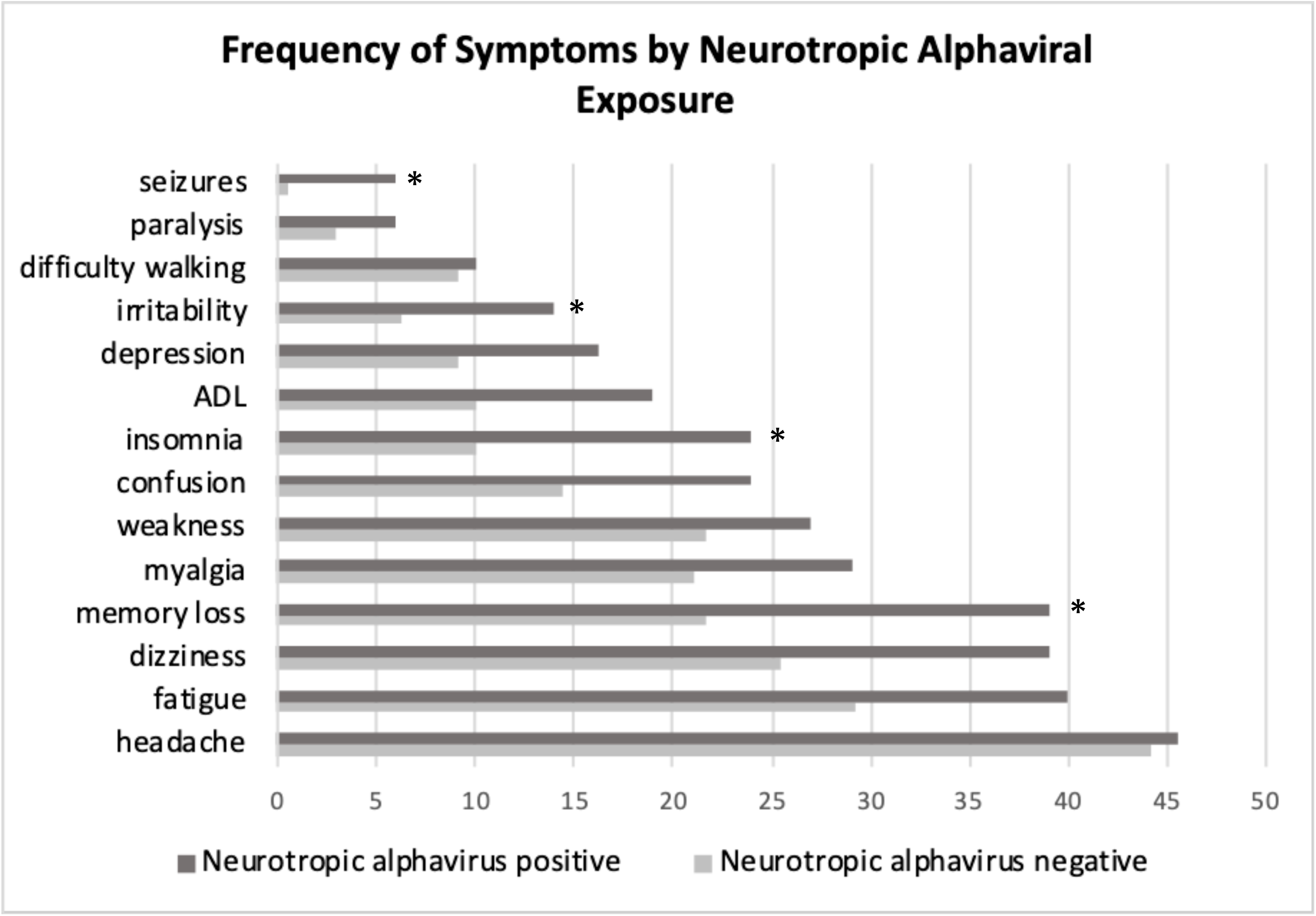
Frequency of self-reported signs and symptoms amongst the 2010 outbreak cohort studied in 2015 and the 2017 Mogue study population, combined (n=308). Significant differences in rates (p<0.05), controlled for age and sex, are denoted with (*)

## Discussion

We provide new clinical and epidemiological findings on human infection with MADV and VEEV. Our seroconversion results suggest that MADV has become endemic, with continued co-circulation of VEEV in eastern Panama. Our data show that some 2010 MADV patients had no detectable MADV antibodies five years after exposure; interestingly, people exposed to MADV in 2010 had higher rates of VEEV seroconversion than those who were originally seronegative for alphaviruses. On the other hand, subjects with prior exposure to VEEV tended to seroconvert to MADV at lower rates that those who were originally alphavirus-seronegative, though this was not statistically significant. These data suggest that MADV exposure leads to enhanced susceptibility against VEEV, but not vice-versa. The mechanism underlying this enhanced susceptibility to VEEV following MADV exposure is unclear. Increased risk of acquiring alphavirus sympatric infections may reflect their similar enzootic habitat and overlapping epidemiological risk of acquisition [5]. However, we have observed differences in MADV and VEEV vector and host usage, as well as different geographic distribution of disease [4,13]. The seemingly increased susceptibility to VEEV conferred by prior exposure to MADV may also be explained by immune interference. Alphavirus vaccine studies in humans and horses have found that administration of eastern equine encephalitis (EEE) and western equine encephalitis (WEE) vaccines prior to live-attenuated VEE vaccination resulted in a diminished VEEV neutralizing antibody response [17,18]. When VEE vaccination was provided prior to EEE/WEE vaccination, VEEV neutralizing antibody response remained intact [17]. To our knowledge, this is the first epidemiological study to demonstrate asymmetric human cross-immunity findings, even though these viruses were identified almost a century ago in the 1930’s[2,10].

In contrast, VEEV infection appears to generate a robust and durable immune response. Not only do titers remain detectable 5 years after exposure, but the low rate of seroconversion to MADV during this period suggests the presence of cross-protective immunity. The alphaviral vaccine literature provides ample evidence of in vivo examples of cross-protection. For example, hamsters inoculated with an attenuated strain of VEEV experienced a 37% reduction in mortality when subsequently exposed to western equine encephalitis virus (WEEV), and a 59% reduction in mortality when inoculated with EEEV [19]. The duration of this cross-protection appears to be at least 3 months [9]. For the more distantly related Mayaro virus, however, cross-protection was shorter lasting [9,20]. While such animal studies show in vivo protection to sequential infection, passive transfer of neutralizing antibodies may not confer protection to heterologous viruses [9,20]. Cross-protection may therefore be attributable to a cellular response or a humoral response mediated by non-neutralizing antibodies. There is indeed evidence for cellular cross-reactivity, as seen in murine studies of capsid-specific cytotoxic T-lymphocytes (CTLs) showing cross-reactivity to many Old-World alphaviruses. The presence of these CTLs did not alter primary viremia, but did enhance viral clearance [21]. While neutralizing antibodies tend to be virus-specific, non-neutralizing antibodies can be cross-reactive in nature [22], and engage in antibody-dependent cell mediated cytotoxicity (e.g. in HIV) [23–25].

Seroreversion was documented in 14.3% of 2010 MADV-positive individuals. It is possible that neutralizing antibody titers wanes over time and dropped below our limits of detection. Whether these individuals are newly at risk for MADV infection remains uncertain. In the case of hepatitis B, for example, it was demonstrated that in the face of declining antibody titers following vaccination, hepatitis B infection rates rose in Senegalese children [26]. While the causes of antibody decay are poorly understood, short-lived humoral immune responses may be due to a failure to effectively induce memory B cells and long-lived plasma cells [27]. Such failure could result from a lack of the pathogen’s immunogenicity, though other mechanisms have been described, such as targeted immunosuppression of germinal centers by Borrelia burgdorferi, the causative agent of Lyme disease [27].

Memory loss, dizziness, fatigue, difficulty concentrating, confusion, depression, irritability, myalgia, insomnia, and impairment of daily living activities were more frequent in subjects exposed to VEEV and/or MADV. After adjusting for sex and age, increased seizures, irritability, insomnia and memory loss remained statistically significant. Seizures and paralysis were mainly observed in severe cases of MADV encephalitis during the 2010 outbreak. However, seizures were not present in any of the 2010 VEE subjects, in contrast to reports of VEE sequelae in children in Colombia [28]. Indeed, on the whole, our 2010-2015 cohort of VEEV-positive subjects did not present with severe cases of neurological disease. This may reflect variations in virulence among VEEV strains [29], though there are historical reports of severe and fatal cases of enzootic VEE in Panama [16].

Seven of nine adults reported fatigue 9 months following a VEEV outbreak in Texas in 1971 [6]. Our results more closely resemble descriptions of sequelae following WEEV infection [30] as well as West Nile virus [31,32]. Memory loss, learning impairments, and behavioral changes are noted in approximately half of patients following acute illness due to neurotropic Alphaviruses [33]. While there does not appear to be any precedent for cognitive testing of subjects exposed to alphaviral infections, there have been several such studies in patients recovered from West Nile virus (WNV). For example, Murray et al [34], noted that at 8 years from initial infection (neuroinvasive and febrile WNV), 40% of patients still reported sequelae. Fatigue, weakness and depression were most common, but 9% of these patients also reported memory loss and confusion. In a companion study, the authors investigated magnetic resonance image (MRI) findings in these patients, and noted significant cortical thinning in multiple regions, including the posterior cingulate cortex and parahippocampal region, compared to normative controls [31]. We suspect that the true burden of MADV and VEEV extends substantially beyond the acute phase of illness. Understanding the full scope of such sequelae is paramount in this region, given the high prevalence of MADV and VEEV exposure.

Interestingly, the majority of MADV- and VEEV-positive individuals in our study did not recall having encephalitis or severe neurological signs/symptoms. Other studies also have suggested that the majority of the encephalitic alphaviral infections present as a self-limited febrile illness[2,35]. Thus, the high rates of self-reported neurological sequelae in this study suggest that long-term neurological sequelae occur even after mild to moderate clinical presentations.

Our study has several limitations. Because the follow-up study was undertaken 5 years after the outbreak, we were not able to determine the timing of seroconversion during the 5-year period between data collection. It is therefore possible that the majority of the seroconversions occurred shortly after the 2010 outbreak. However, we did note that one of the subjects presented with MADV IgM in 2015, suggesting recent infection. In addition, our sample size was limited because our cohort was generated from subjects originally tested during an outbreak and there were many subjects that could not be traced. Therefore, our data may not be generalizable. Furthermore, there may have been bias in reporting neurological signs and symptoms stemming from the fact that the subjects were selected based on their inclusion in the 2010 outbreak studies. We sought to counteract this by including neurological symptom data from a separate, cross-sectional study conducted in 2017.

In summary, our results demonstrate that MADV remains in circulation and is an important human pathogen in Panama. We further describe other novel findings, such as the decay in MADV antibody in some individuals, the possibility of cross-protective immunity conferred by VEEV but not by MADV, and increased susceptibility to VEEV conferred by prior MADV exposure. These findings have implications for vaccine development and merit further study. In addition, we provide preliminary information on the persistence of neurologic symptoms following MADV and VEEV infection. Future investigation into the duration and magnitude of such sequelae, as well as underlying mechanisms and risk factors, may benefit not only those affected by these viruses, but also the larger population exposed similar neurotropic arboviruses.

## Data Availability

All the information is included in the manuscript. The authors may be contacted at Gorgas Memorial Institute (GMI) for additional information. All information is included in the manuscript. Our epidemiological data cannot be made publicly available for legal reasons (e.g., public availability would compromise patient privacy) by Law 68, 2003 (http://200.46. 254.138/APPS/LEGISPAN/PDF_NORMAS/2000/ 2003/2003_531_2443.PDF).

## Acknowledgments

We are deeply grateful to Mr. Abadia from the Ministry of Health at Taimatí, Darien, for help with tracing subjects and navigating the waters of the Panamanian Pacific coast to far flung villages. We also thank Dr. Sandra Lopez for helping us deliver laboratory results to the cohort subjects, and Jorge Maguiña for help with database maintenance.

## Disclaimers

The opinions expressed by authors contributing to this work do not necessary reflect the opinions of the Gorgas Memorial Institute of Health Studies, The Panamanian Government or the institutions with which the authors are affiliated.

